# Mindsets Predict Physical Activity and Relate to Chosen Management Strategies in Individuals with Knee Osteoarthritis

**DOI:** 10.1101/2021.03.31.21254737

**Authors:** Melissa A. Boswell, Kris M. Evans, Sean R. Zion, Danielle Z. Boles, Jennifer L. Hicks, Scott L. Delp, Alia J. Crum

## Abstract

**Objectives:** We compared mindsets about physical activity among those with and without knee osteoarthritis and investigated if these mindsets relate to physical activity level and symptom management.

**Methods:** Participants with (n=150) and without (n=152) knee osteoarthritis completed an online survey at study enrollment (T1). Participants with knee osteoarthritis repeated the survey three weeks later (T2; n=62). The mindset questionnaire, scored from 1-4, assessed the extent to which individuals associate the process of exercising with less appeal-focused qualities (e.g., boring, painful, isolating, and depriving) vs. appeal-focused (e.g., fun, pleasurable, social, and indulgent) versus. Using linear regression, we examined the relationship between mindset and having knee osteoarthritis, and, in the subgroup of participants with knee osteoarthritis, the relationship between mindset at T1 and physical activity (via the Physical Activity Scale for the Elderly) at T2. We also compared mindsets between those who use medication for management and those who use exercise.

**Results:** A less appeal-focused mindset regarding physical activity was marginally associated with having knee osteoarthritis (β=-0.125, *P*=0.096). Within the knee osteoarthritis group, a more appeal-focused mindset predicted higher future physical activity (β=20.68, *P*=0.039), controlling for current physical activity, demographics, and health. Individuals that used exercise with or without pain medication or injections had more appeal-focused process mindsets than those who used medication or injections without exercise (*P*<0.001). Further, the process mindset inventory demonstrated strong internal consistency (α=0.92 at T1 for n=150 and α=0.92 at T2 for n=62) and test-retest reliability (ICC>0.841, *P*<0.001) within the knee osteoarthritis population.

**Conclusion:** In individuals with knee osteoarthritis, mindsets predict future physical activity levels and relate to an individual’s management strategy. Mindsets are a reliable and malleable construct and may be a valuable target for increasing physical activity and improving adherence to rehabilitation strategies involving exercise among individuals with knee osteoarthritis.

## Introduction

There are an estimated 14 million individuals in the United States with knee osteoarthritis[1], a disease characterized by soft tissue degradation in the knee joint, namely the cartilage and meniscus. This incidence has grown as the population has aged and obesity has become more prevalent[1]. Knee osteoarthritis is a leading cause of disability and is often associated with progressively worsening pain and dysfunction due to a lack of disease-modifying therapies[2].

Long-term engagement in physical activity can improve pain and function[3] and is related to better cartilage health[4] in individuals with knee osteoarthritis. As an additional benefit, physical activity helps prevents loss of muscle strength, which contributes to disability[5]. Despite these benefits, management of knee osteoarthritis with physical activity is under-utilized[6], and long-term adherence to physical activity programs within the knee osteoarthritis population is low[7]. Limited engagement in physical activity makes individuals with knee osteoarthritis more susceptible to functional decline[8].

Emerging research has highlighted the powerful influence of mindsets about physical activity on engagement in physical activity. Mindsets are core assumptions about a domain or category which orient individuals to a particular set of attributions, expectations, and goals (a “meaning system”)[9]. Mindsets have been studied in a variety of domains, with one of the most well-studied being education[10] (e.g., mindsets about intelligence as “fixed” or “malleable”). Recent studies have investigated mindsets about health-related constructs, including stress[9], illness[11], and physical activity[12]. For example, an improvement in an individual’s mindset about the adequacy of their physical activity (i.e., my activity level is adequate and thus beneficial to my health) significantly decreased weight, blood pressure, and body fat compared to a control group, and without an increase in activity[13]. The ability of mindset to influence one’s health and well-being, in addition to behavior, differentiates it from other well-studied constructs related to physical activity levels. For example, an adaptive adequacy mindset predicts greater self-efficacy and physical activity levels and, separately, predicts better perceived health[14].

Another mindset about physical activity regards the *process* of being physically active. These mindsets are defined as the extent to which individuals associate the process of engaging in exercise behaviors with less appeal-focused qualities (e.g., difficult, unpleasant, stressful, inconvenient, boring, isolating, and depriving)[15] versus more appeal-focused (e.g., easy, pleasurable, relaxing, convenient, fun, social, and indulgent). Research on healthy individuals has shown that mindsets about the appeal of physical activity can be improved and are associated with health status and predict physical activity involvement. For example, a brief intervention that emphasized the social, fun, and self-indulgent aspects of exercise shifted individuals’ mindsets about the process of physical activity to be more appeal-focused (referred to as an “appeal-focused mindset” for brevity) and increased exercise adherence in a 10-week fitness class and motivation for future exercise[15]. Further, these mindsets predicted self-reported physical activity, controlling for perceived importance of health and self-efficacy[15]. Appeal-focused mindsets are theorized to be more adaptive because they foster more intrinsically motivating[16] and enjoyable[17] experiences with physical activity.

Individuals with knee osteoarthritis may view physical activity as less appeal-focused than the general population due to the unique challenges of joint pain, swelling, and stiffness[3]; functional limitations[3]; and misconceptions about physical activity as it relates to their osteoarthritis status[7,18]. For example, a commonly held view of individuals with osteoarthritis is that osteoarthritis is a disease of “wear and tear”[18]. These beliefs and misconceptions, along with one’s experiences and social interactions, inform an individual’s mindset. The mindsets that individuals with knee osteoarthritis hold about physical activity may be particularly impactful because of their associated meaning system made up of mindset-congruent attributions (e.g., “physical activity is unpleasant because it’s bad for my knees” vs. “physical activity is pleasant because it strengthens my body and improves my joint function”), expectations (e.g., “physical activity is boring and will make me tired and achy” vs. “physical activity is fun and will make me energized and refreshed”), and goals (avoid physical activity vs. seek out ways to adapt physical activity and use it as a means for rehabilitation). In this way, mindsets about physical activity may affect physical activity participation and management strategy preference in the knee osteoarthritis population beyond other known determinants of physical activity, including factors such as age[19], gender[19], body mass index (BMI)[20], overall health[21], and pain due to knee osteoarthritis [20]. Mindsets have not been evaluated in the knee osteoarthritis population. Yet, understanding and intervening to improve mindsets about the appeal of physical activity in the knee osteoarthritis population may increase physical activity in people with knee osteoarthritis.

This study examined the mindsets that individuals with knee osteoarthritis hold about physical activity with the process mindset inventory. We hypothesized that individuals with knee osteoarthritis would have less appeal-focused mindsets about physical activity than those without knee osteoarthritis. Additionally, we hypothesized that more appeal-focused mindsets would predict future physical activity levels and relate to increased use of exercise as an individual’s knee osteoarthritis management strategy. As a secondary analysis, we assessed the reliability and internal consistency of the process mindset inventory among individuals with knee osteoarthritis.

## Patients and Methods

### Participants

We recruited individuals with a self-reported clinical diagnosis of knee osteoarthritis and individuals without knee osteoarthritis within the United States using Centiment Research, an online survey platform. We included participants in the study if they were between the ages of 45-85 years. We excluded participants who had a previous total knee arthroplasty surgery or other knee surgery, could not speak English, or did not complete the survey. The online survey was completed by 150 individuals with knee osteoarthritis and 152 individuals without knee osteoarthritis at time point 1 (T1). We asked the group with knee osteoarthritis to repeat the survey three weeks later at time point 2 (T2). A subset of these participants (62 of the 150 participants; 41%) repeated the survey. There were no significant differences in any of the variables measured between those who completed the survey at T2 and those who did not. We obtained approval for the study from the Stanford University Institutional Review Board and digital informed consent from all participants.

### Measures

#### Physical activity

We assessed physical activity levels using the Physical Activity Scale for the Elderly (PASE). The PASE includes the frequency of light, moderate, and strenuous work and leisure activities and is a validated measure of self-reported physical activity for individuals with osteoarthritis[22].

#### Process of physical activity mindset

To assess mindsets about physical activity, we used the process mindset inventory (Table S1). The process mindset inventory is a one-factor scale developed and validated by Boles and colleagues[15] to assess mindsets about the process of engaging in physical activity (e.g., physical activity is difficult/easy, unpleasant/pleasurable, boring/fun). The scale is 7 items, measured on a 4-point scale, and scored from 1-4, with higher scores reflecting a more appeal-focused mindset about physical activity.

#### Management strategy

We determined individuals’ knee osteoarthritis management strategies with the open-ended response question, “In your own words, describe how you manage and/or improve the symptoms of osteoarthritis.”

#### Health

We assessed overall physical and mental health status using the validated PROMIS v.1.1 Global Health Short Form[23]. The Global Health Short Form is a ten-item survey that measures overall physical function, fatigue, pain, emotional distress, and social health in healthy and clinical adult populations.

#### Knee pain and function

We captured osteoarthritis-related knee pain and functioning using the likert version of the Western Ontario and McMaster Universities Osteoarthritis Index (WOMAC)[24]. The WOMAC is a validated and reliable disease-specific 24-item measure of knee pain, stiffness, and function for patients with knee osteoarthritis [24].

Other information collected included gender, age, BMI calculated from height and weight, health status, and knee pain and functioning. The survey for participants with knee osteoarthritis included all described assessments; the survey for participants without knee osteoarthritis included all described assessments except the WOMAC.

### Statistical analyses

We used R (v3.5.0)[25] for analyses and used R package ggplot2[26] to produce figures. In all regressions, we standardized the continuous independent variables and examined the coefficients (β) with 95% confidence intervals (CI) and the adjusted coefficient of determination (R^2^). The a priori level of significance, α, for all statistical tests was 0.05.

We assessed the internal consistency and test-retest reliability of the process mindset within the group of participants with knee osteoarthritis to measure its reliability in the knee osteoarthritis population. We calculated Cronbach’s alpha[27] to assess the internal consistency of the process mindset at baseline and follow-up (with R package psych[28]). We calculated the intraclass correlation coefficient to assess test-retest reliability using a two-way mixed-effects model with absolute agreement between the mean process mindset score at baseline and follow-up (with R package irr[29]).

To test for differences in physical activity level, demographic variables, and health between the knee osteoarthritis and control populations, we calculated the standardized mean difference (SMD)[30] (with R package stddiff[31]). We chose an SMD of less than 0.1 to indicate a negligible difference, a threshold recommended to determine imbalance[32].

We used multivariate linear regression modeling (with R package lmSupport[33]) to determine whether having knee osteoarthritis was a predictor of the process mindset when controlling for other factors that may influence mindset. The dependent variable was the process mindset at T1. The independent variables were a binary (0/1) for the presence of knee osteoarthritis (with 1 indicating knee osteoarthritis), a binary for gender (with 1 indicating female), age, BMI, global health at T1, and physical activity level at T1.

To test whether the process mindset predicted future physical activity levels in individuals with knee osteoarthritis, we used a multivariate linear regression with the PASE measure at T2 as the dependent variable and the process mindset at T1 as the independent variable. We controlled for demographics, health, knee pain and functioning, and physical activity level at T1 by adding all of these as independent variables in the model. In an exploratory analysis, we evaluated the relationship between physical activity level at T2 and each item of the process mindset inventory at T1.

Three researchers (MB, KE, and a member of the lab of AC) reviewed the open-ended management question responses to determine a set of strategy categories (e.g., pain medication or injections, independent physical activity, and diet or weight management). Once a set of categories was agreed upon, two researchers (MB and a member of the lab of SD) separately coded all 150 responses for whether the strategy category was apparent. We calculated inter-rater reliability for the two most common themes. Cohen’s kappa was 0.987 (P < 0.001) and 0.931 (P < 0.001) for medication or injections and exercise, respectively, indicating “almost perfect” agreement[34]. A third researcher (another member of the lab of SD) determined the final coding decision for all disagreements between the two coders. We used independent t-tests to assess differences in mindset between those who manage their osteoarthritis (1) with medication or injections but without exercise, (2) with exercise but without medication or injections, and (3) with both exercise and medication or injections.

## Results

### Internal consistency and test-retest reliability

The process mindset inventory demonstrated strong internal consistency (α = 0.92 with mean (SD) = 2.25 (0.69) at T1 for n = 150; α = 0.92 with mean (SD) = 2.24 (0.65) at T2 for n = 62) and test-retest reliability (ICC > 0.841, 95% Confidence Interval (CI) = [0.749,0.901], *P* < 0.001) within the knee osteoarthritis population.

### Between group differences

The group with knee osteoarthritis had an older age (*P* = 0.006), a higher BMI (P < 0.001), lower global health (*P* < 0.001), and lower levels of physical activity (*P* < 0.001) than individuals without knee osteoarthritis (Table 1). There were no differences in gender distribution between the two groups (*P* = 0.640).

**Table 1.**
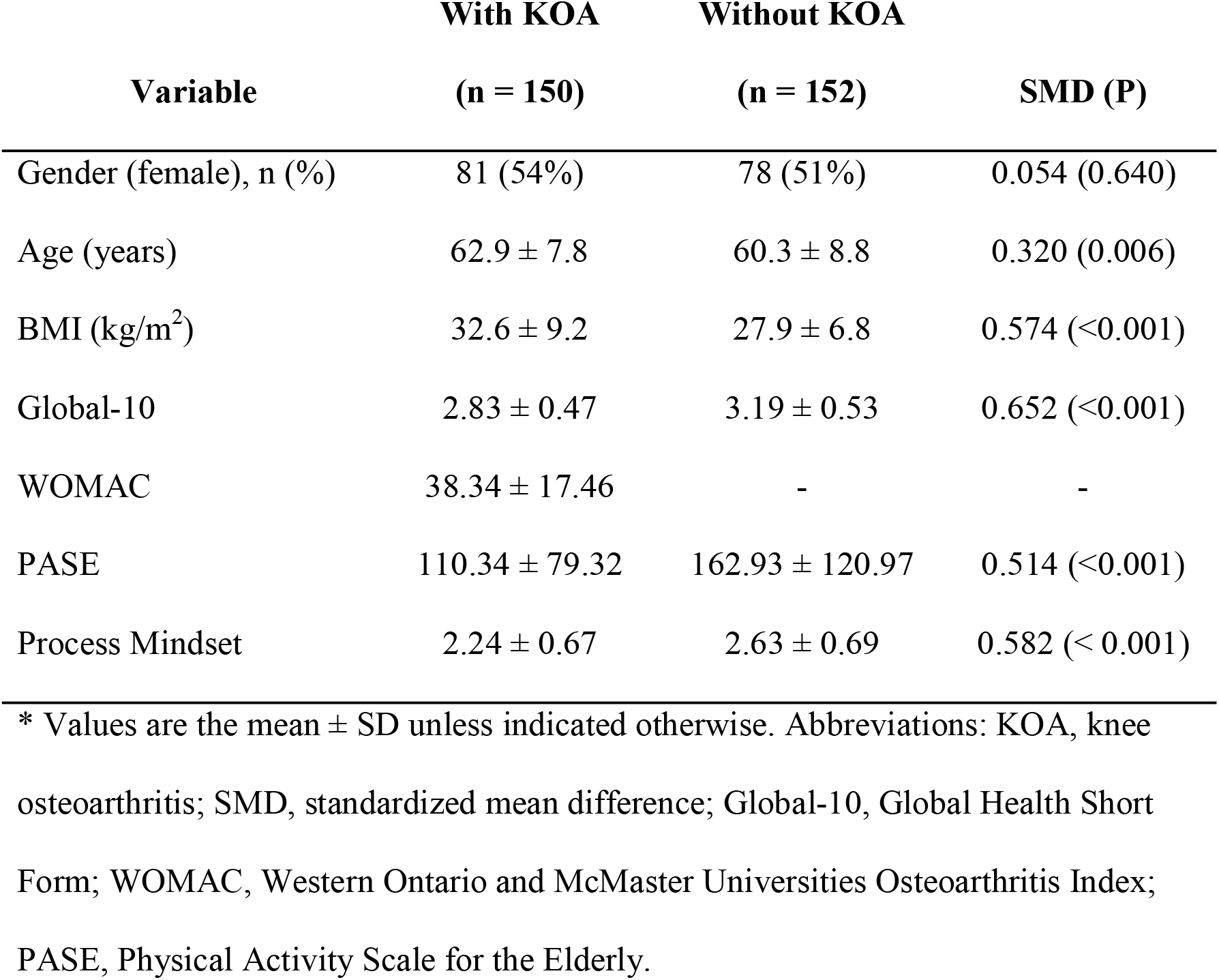
Characteristics of participants and standardized mean difference (SMD) between participants with and without knee osteoarthritis (KOA) at time point 1 (T1). Individuals with knee osteoarthritis were older and had higher BMI, lower global health, and lower physical activity levels than individuals without knee osteoarthritis.

| Variable | With KOA<br>(n = 150) | Without KOA<br>(n = 152) | SMD (P) |
| --- | --- | --- | --- |
| Gender (female), n (%) | 81 (54%) | 78 (51%) | 0.054 (0.640) |
| Age (years) | 62.9 ± 7.8 | 60.3 ± 8.8 | 0.320 (0.006) |
| BMI (kg/m<sup>2</sup>) | 32.6 ± 9.2 | 27.9 ± 6.8 | 0.574 (<0.001) |
| Global-10 | 2.83 ± 0.47 | 3.19 ± 0.53 | 0.652 (<0.001) |
| WOMAC | 38.34 ± 17.46 | - | - |
| PASE | 110.34 ± 79.32 | 162.93 ± 120.97 | 0.514 (<0.001) |
| Process Mindset | 2.24 ± 0.67 | 2.63 ± 0.69 | 0.582 (< 0.001) |

\* Values are the mean ± SD unless indicated otherwise. Abbreviations: KOA, knee osteoarthritis; SMD, standardized mean difference; Global-10, Global Health Short Form; WOMAC, Western Ontario and McMaster Universities Osteoarthritis Index; PASE, Physical Activity Scale for the Elderly.

### Predictiveness of having knee osteoarthritis on mindset

Having knee osteoarthritis was marginally associated with a less appeal-focused mindset by 0.125-points (CI = [-0.273, 0.022], *P* = 0.096) when controlling for gender, age, BMI, global health, and physical activity level (Table 2). A lower score corresponds to a mindset that physical activity is less appeal-focused (e.g., more boring, isolating, and depriving). For reference, a previous study found that a 0.11-point higher process mindset score led to higher attendance during a 10-week fitness class and increased the likelihood of enrolling in another physical education class in the following quarter[15]. Additional factors related to a less appeal-focused process mindset were gender (being female; β = -0.137, CI = [-0.271, -0.002, *P* = 0.047]), lower global health (β = 0.144, CI = [0.066, 0.222], *P* < 0.001), lower physical activity levels (β = 0.247, CI = [0.172, 0.321], *P* < 0.001), and, marginally, a higher BMI (β = -0.071, CI = [-0.144, 0.001], *P* = 0.053).

**Table 2.** Linear regression on the process mindset for participants with and without knee osteoarthritis (n = 302). Coefficients (β) are presented with a 95% confidence interval (CI). Predictors of a less appeal-focused process mindset were gender (being female), lower global health, and lower physical activity levels. A higher BMI and having knee osteoarthritis may be marginally predictive of a less appeal-focused mindset.

| Dependent Variable | Independent Variables | $\beta$ (95% CI) | P | Adj. R<sup>2</sup> | F |
| --- | --- | --- | --- | --- | --- |
| Process Mindset | Gender | -0.137 (-0.271, -0.002) | 0.047 | 0.303 | 22.8<br>(p < 0.001) |
|  | Age | -0.040 (-0.111, 0.030) | 0.262 |  |  |
|  | BMI | -0.071 (-0.144, 0.001) | 0.053 |  |  |
|  | Global-10 | 0.144 (0.066, 0.222) | <0.001 |  |  |
|  | PASE | 0.247 (0.172, 0.321) | <0.001 |  |  |
|  | **Knee osteoarthritis (binary)** | **-0.125 (-0.273, 0.022)** | **0.096** |  |  |

Abbreviations: Global-10, Global Health Short Form; PASE, Physical Activity Scale for the Elderly.

### Predictiveness of mindset for future physical activity

The process mindset at T1 predicted future physical activity level (β = 20.68, CI = [1.06, 40.30], *P* = 0.039) while controlling for demographics, health, knee pain and functioning, and physical activity level at T1 (Table 3). This result means that a one standard deviation increase in the process mindset increases the PASE score by almost 21 points. A 21-point increase in the PASE corresponds to, for example, going from performing a physical activity “seldom” for 1-2 hours per day to “often” for 1-2 hours per day (quoted words are from the PASE scale). Further, the process mindset was the only variable aside from the PASE at T1 that was predictive of physical activity level at T2. An additional sensitivity analysis with the same model, but removing an outlier with a PASE score at T2 greater than three standard deviations above the mean reduced this estimated effect to 12-points (CI = [-2.38, 26.40], *P* = 0.100).

**Table 3.** Linear regression on physical activity level (PASE) at time point 2 (T2) for participants with knee osteoarthritis (n = 62). A more appeal-focused process mindset and higher physical activity levels at T1 were predictive of higher physical activity levels at T2.

| Dependent Variable | Independent Variables | $\beta$ (95% CI) | P | Adj. R<sup>2</sup> | F |
| --- | --- | --- | --- | --- | --- |
| PASE (T2) | Age | -3.16 (-21.48,15.15) | 0.730 | 0.49 | 9.262<br>(p < 0.001) |
|  | Gender | -26.21 (-60.55, 8.12) | 0.132 |  |  |
|  | BMI | -6.77 (-24.99, 11.46) | 0.460 |  |  |
|  | Global-10 (T1) | 14.03 (-8.62, 36.67) | 0.220 |  |  |
|  | WOMAC (T1) | 11.16 (-10.20, 32.52) | 0.300 |  |  |
|  | PASE (T1) | 47.52 (28.55, 65.95) | <0.001 |  |  |
|  | **Process Mindset (T1)** | **20.68 (1.06, 40.30)** | **0.039** |  |  |

Abbreviations: Global-10, Global Health Short Form; WOMAC, Western Ontario and McMaster
Universities Osteoarthritis Index; PASE, Physical Activity Scale for the Elderly.

### Individual mindset inventory item correlations

Additional exploratory analyses revealed correlations between the individual process mindset items at T1 and future activity levels (Table S2). Out of these items, ranking the process of physical activity as easier (on a scale of difficult to easy) had the strongest correlation (r = 0.52, *P* < 0.001), whereas ranking the process of physical activity as more fun (on the scale of boring to fun) had the weakest correlation (r = 0.28, *P* = 0.029).

### Relation between management strategy and mindset

We determined nine distinct management strategies from responses to the open-ended question about knee osteoarthritis symptom management (Table 4). Almost 50% of responses mention pain medications or injections (n = 74), while close to 27% of responses mention physical activity (n = 41). Additional strategies included self-soothing (n = 25), nothing (n = 22), impose physical limitations (n = 21), home remedies (n = 11), rest (n = 9), talk to a doctor (n = 6), diet or weight management (n = 6), supervised physical therapy (n = 5), and self-motivation (n = 4). Individuals that used exercise with or without pain medication or injections had more appeal-focused process mindsets than those who used medication or injections without exercise (t = -2.95, df = 13.21, *P* = 0.011, CI = [-1.08, -0.17], mean = (2.02, 2.65) and t = -5.63, df = 64.80, *P* < 0.001, CI = [-0.95, -0.45], mean = (2.02, 2.72), respectively; Figure 1). Mindsets did not differ between individuals who used physical activity with medication or injections and those who used physical activity without medication or injections (t = 0.34, df = 15.13, *P* = 0.74, CI = [-0.39, 0.54], mean = (2.72, 2.64)).

**Table 4.** The apparent themes from responses to the open-ended question, “In your own words, describe how you manage and/or improve the symptoms of osteoarthritis.” Almost 50% of responses mentioned pain medications or injections, while close to 27% of responses mention physical activity (n = 150).

| Theme | #/150 | Example |
| --- | --- | --- |
| Pain Medications or Injections | 74 | Pain medication every day; Cortisone shots |
| Independent Physical Activity | 41 | Exercise daily in all kinds of weather |
| Self-Soothing | 25 | Hot compress; Muscle rub; Creams and gels |
| Nothing | 22 | Just put up with it; Just continue on with normal activities |
| Impose Physical Limitations | 21 | Go very slowly & think before I make any sudden or stretching movements |
| Home Remedies | 11 | I drink apple cider vinegar; Soaking my knees with Epsom salts |
| Rest | 9 | Rest when my body tells me to |
| Doctor | 6 | Have switched doctors hoping to get relief from all the pain. |
| Diet or Weight Management | 6 | Clean eating; Giving up diet soda |
| Supervised Physical Therapy | 5 | Physical therapy every day getting stronger |
| Self-Motivation | 4 | I am motivated to move beyond the pain; Mind over matter |

**Figure 1.**
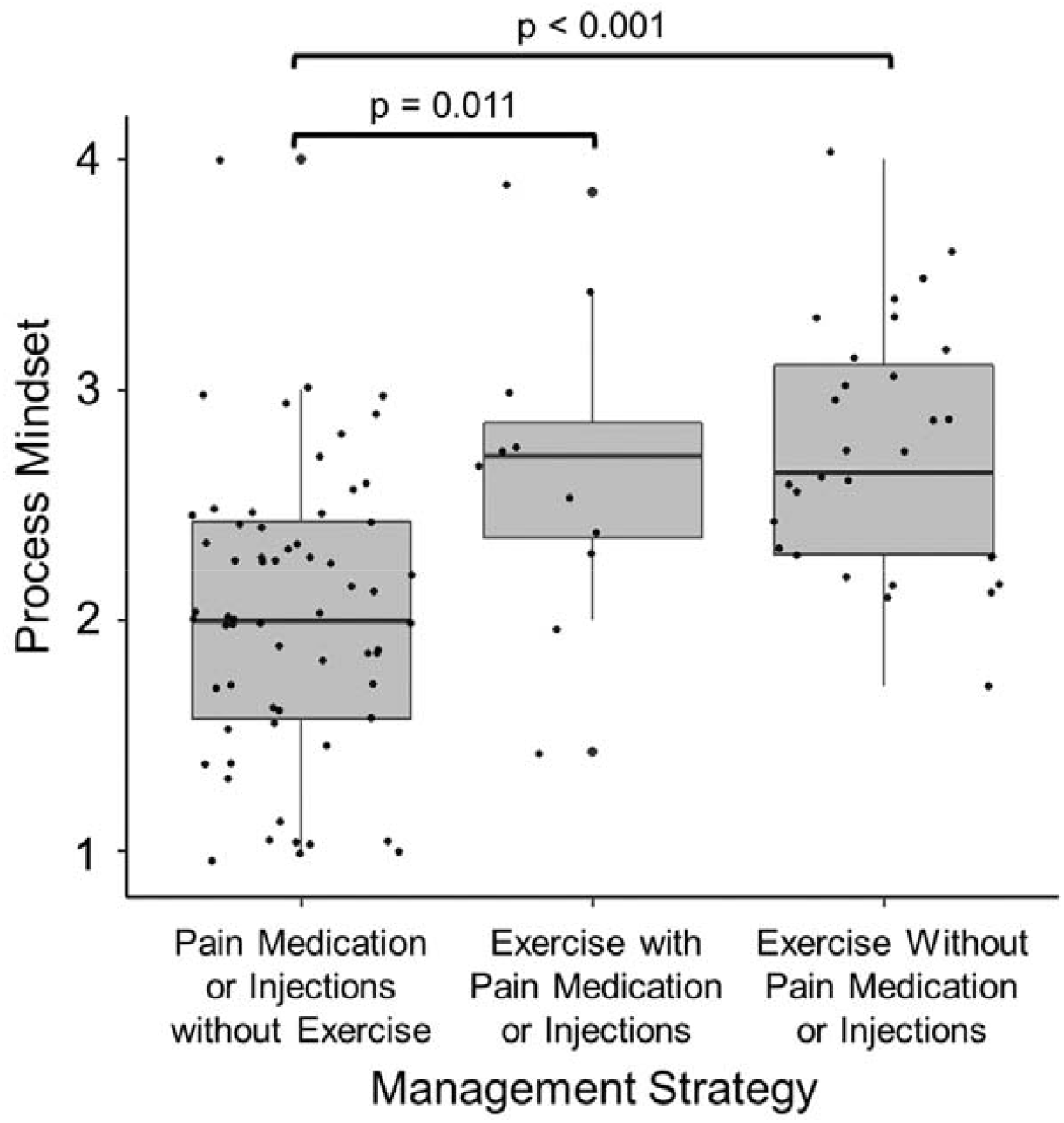
Differences in process mindset based on knee osteoarthritis management strategy. Those who use exercise as a management strategy in addition to or without pain medication or injections had a significantly more adaptive mindset about the process of physical activity than those who use pain medication or injections without exercise (*P* = 0.011 and *P* < 0.001, respectively).

## Discussion

We found that individuals with knee osteoarthritis had marginally less appeal-focused mindsets about physical activity than individuals without knee osteoarthritis while controlling for demographic (e.g., gender, age) and health (e.g., BMI, global health) variables and physical activity level. Within the sample of participants with knee osteoarthritis, mindsets about physical activity predicted future physical activity level, above and beyond other known determinants of physical activity, including demographic and health variables, knee pain and functioning, and current physical activity level. Although a sensitivity analysis revealed this relationship was only marginal, this is likely due to a smaller than sufficient sample to detect a small effect size. Finally, individuals who chose exercise as a strategy for osteoarthritis management had a more appeal-focused mindset about physical activity than those who chose pain medications or injections without exercise.

Having knee osteoarthritis was only marginally associated with a less appeal-focused process mindset, likely due to the effects of confounding variables. Those with knee osteoarthritis had a higher BMI, lower global health, and lower physical activity levels than those without knee osteoarthritis. These differences are consistent with previous findings that show associations between knee osteoarthritis prevalence and higher weight[35], increased physical and mental health challenges[3], and lower levels of physical activity[36]. Note that the difference from the linear regression analysis is smaller than the raw difference in means between the group with knee osteoarthritis and the group without knee osteoarthritis (mean ± standard deviation: 2.24 ± 0.67 vs. 2.63 ± 0.69, respectively; SMD = 0.582, *P* < 0.001), highlighting the importance of controlling for differences in BMI, health, and other factors when comparing individuals with and without knee osteoarthritis. While joint pain, functional limitations, and maladaptive misconceptions about knee osteoarthritis may negatively impact the mindset of an individual with knee osteoarthritis, the mindset scores in this study display a range of mindsets about physical activity. Some individuals indeed hold a more appeal-focused mindset, demonstrating that a less appeal-focused mindset is not inevitable in individuals with knee osteoarthritis, and supporting the potential for improving this mindset.

The 7-question process mindset scale can be efficiently and reliably administered in the knee osteoarthritis population, suggesting feasibility for use in more extensive studies. Capturing personal beliefs and expectations about physical activity is often done via qualitative surveys[21,37]. In contrast, the process mindset captures beliefs and expectations about physical activity quantitatively and facilitates analysis of the efficacy of potential interventions. Additionally, whereas many studies and interventions target a particular belief or expectation about specific health goals (e.g., “Running 3 days per week for the next 3 months is: *Not enjoyable*… *Enjoyable*”)[17], this study sought a more general mindset approach targeting the experience of being physically active (e.g., “The process of physical activity is: *Stressful… Relaxing*”). This general approach is valuable for intervening more broadly on the wide range of physical activities one might perform, which may be more likely to be adopted for long-term physical activity engagement.

These findings can help guide clinicians’ strategies for increasing physical activity participation and adherence to rehabilitative programs involving exercise in patients with knee osteoarthritis by improving mindsets about the process of physical activity. For example, a clinician might help patients with knee osteoarthritis adapt their current type or duration of physical activity to feel “easier” or more achievable, rather than simply suggesting the knee osteoarthritis activity guidelines of 30 minutes of moderate-intensity physical activity for three days per week[38]. Another strategy to improve the process mindset may include helping individuals think creatively about different types of physical activity they may enjoy (e.g., yoga, swimming, gardening, dancing, or walking the dog) while highlighting how it can also be social (e.g., walking with a friend, playing with grandchildren, or joining group exercise classes). The time of diagnosis may be a particularly important opportunity to shift mindsets about physical activity. For example, one could suggest a newly diagnosed patient try various low-to moderate-intensity activities while focusing on what they enjoy about the activity.

Our study had several limitations. First, while all participants with knee osteoarthritis indicated a previous clinical diagnosis, we did not obtain radiographic confirmation of knee osteoarthritis. Additional limitations are a potential response bias towards those with internet access and self-selection bias. While we did not evaluate these biases, the survey was available nationally and had characteristics such as gender, race, income, and education levels similar to the general U.S. population. Still, we did not weight the data to obtain a nationally representative sample, which may lead to differences from the general population. Further, the loss to follow-up may have introduced bias through unmeasured factors. Another limitation is that we did not collect objective measures of physical activity. The PASE is validated and widely used for this population; however, self-reported physical activity may be influenced by one’s mindset beyond objective physical activity and remains a question for future investigation. Future studies may benefit from the inclusion of objective physical activity and health outcomes. Future studies should also include a participant sample large enough to detect small effect sizes.

In summary, we surveyed mindsets about the process of physical activity in individuals with and without knee osteoarthritis and assessed the extent to which these mindsets are related to physical activity participation. Results show that within the knee osteoarthritis population, mindset predicted future physical activity when controlling for other factors that commonly influence physical activity levels and related to an individual’s preferred symptom management strategy. Our findings suggest that improving mindsets about physical activity in the knee osteoarthritis population may increase physical activity participation, and as a result, improve health and osteoarthritis outcomes. Future research should identify effective strategies to deliver mindset interventions to individuals with knee osteoarthritis and measure if they indeed change mindsets, physical activity, and health.

## Data Availability

The data is not currently available for public access. Please contact the corresponding author with additional questions.

## Supplemental Tables

**Table S1.** The process mindset inventory and accompanying instructions. A four-point scale should be used for all seven statements: 1 = Very “bad”; 2 = Somewhat “bad”; 3 = Somewhat “good”; 4 = Very “good”. The extent to which a participant endorses an appeal-focused mindset about the process of physical activity can be obtained by calculating the mean score for all items. A higher score indicates greater agreement with an appeal-focused mindset.

**Exercising is:**
|  |  |  |  |
| --- | --- | --- | --- |
| Very difficult | Somewhat difficult | Somewhat easy | Very easy |
| Very unpleasant | Somewhat unpleasant | Somewhat pleasurable | Very pleasurable |
| Very stressful | Somewhat stressful | Somewhat relaxing | Very relaxing |
| Very inconvenient | Somewhat inconvenient | Somewhat convenient | Very convenient |
| Very boring | Somewhat boring | Somewhat fun | Very fun |
| Very lonely | Somewhat lonely | Somewhat social | Very social |
| Very depriving | Somewhat depriving | Somewhat indulgent | Very indulgent |

**Table S2.**
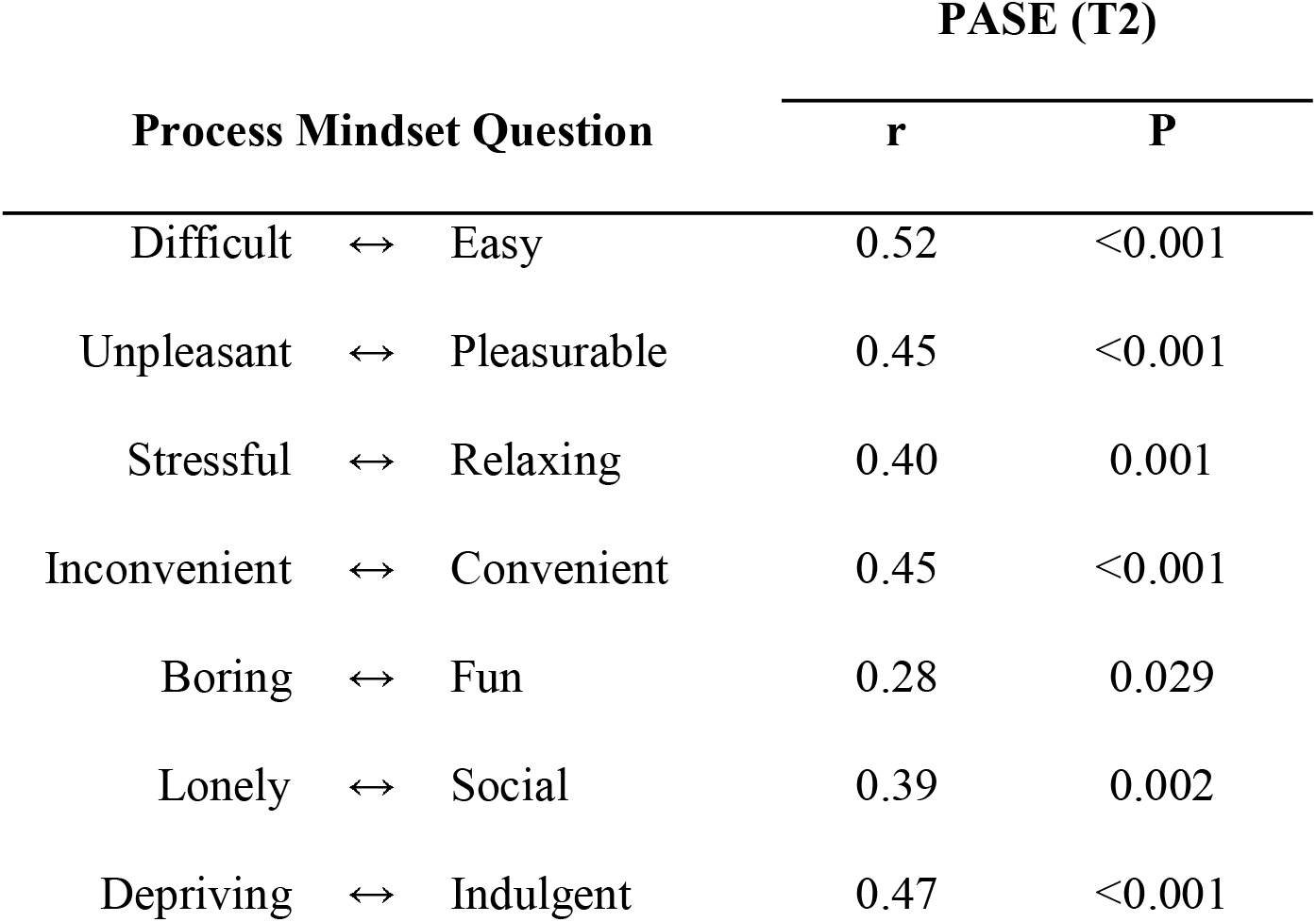
Pearson correlations between physical activity level (PASE) at time point 2 (T2) and each question of the process mindset measure at T1 for participants with knee osteoarthritis (n = 62). All questions were prefaced with “The process of physical activity is…”

